# Early administration of lopinavir/ritonavir plus hydroxychloroquine does not alter the clinical course of SARS-COV-2 infection: a retrospective cohort study

**DOI:** 10.1101/2020.06.05.20123299

**Authors:** Andrea Giacomelli, Gabriele Pagani, Anna Lisa Ridolfo, Letizia Oreni, Federico Conti, Laura Pezzati, Lucia Bradanini, Giacomo Casalini, Cinzia Bassoli, Valentina Morena, Simone Passerini, Giuliano Rizzardini, Chiara Cogliati, Elisa Ceriani, Riccardo Colombo, Stefano Rusconi, Cristina Gervasoni, Dario Cattaneo, Spinello Antinori, Massimo Galli

## Abstract

As it has been shown that lopinavir (LPV) and hydroxychloroquine (HCQ) have *in vitro* activity against coronaviruses, they were used to treat COVID-19 during the first wave of the epidemic in Lombardy, Italy.

The aim of this retrospective intent-to-treat analysis of the hospitalized patients who started off-label treatment with LPV/ritonavir (LPV/r)+HCQ between 21 February and 20 March 2020 was to compare the rate of clinical improvement between those who started the treatment within five days of symptom onset (early treatment, ET) and those who started later (delayed treatment, DT). The association between the timing of treatment and the probability of 30-day mortality was also assessed using uni- and multivariable logistic models.

The study involved 172 patients: 43 (25%) in the ET and 129 (75%) in the DT group. The rate of clinical improvement increased over time to 73.3% on day 30, without any significant difference between the two groups (Gray’s test *P* = 0.213). After adjusting for potentially relevant clinical variables, there was no significant association between the timing of the start of treatment and the probability of 30-day mortality (adjusted odds ratio [aOR] ET *vs* DT = 1.45, 95% confidence interval 0.50-4.19). Eight percent of the patients discontinued the treatment because of severe gastrointestinal disorders attributable to LPV/r.

The timing of the start of LPV/r+HCQ treatment does not seem to affect the clinical course of hospitalised patients with COVID-19. Together with the severe adverse events attributable to LPV/r, this raises concerns about the benefit of using this combination to treat COVID-19.

## 1. INTRODUCTION

The current coronavirus disease 2019 (COVID-19) pandemic caused by severe acute respiratory syndrome-coronavirus-2 (SARS-CoV-2) has seriously affected the public health systems of many countries worldwide (3,917,366 cases and 274,361 deaths as of 10 May 2020) [1].

Although most SARS-CoV-2 infections are self-limiting, about 15% of infected adults develop severe pneumonia requiring supplementary oxygen treatment, and 5% progress to critical illness requiring intensive care [2, 3]. The pathogenetic mechanisms underlying COVID-19 are still not fully understood, but increasing evidence indicates that the clinical deterioration observed during SARS-CoV-2 infection is attributable to direct viral damage followed by virus-induced immune-mediated injury [4]. The rapid spread and severity of COVID-19 has prompted clinicians to identify possible therapeutic strategies on the basis of experimental data or clinical experiences with other coronaviruses such as severe acute respiratory syndrome (SARS) and Middle Eastern respiratory syndrome (MERS).

In late February 2020, Italy was the first Western country to be hit by the COVID-19 epidemic, with the Lombardy region alone recording 81,871 cases and 15,054 deaths as of 11 May 2020 [5]. During the first weeks of the epidemic, a *vademecum* was provided by the Lombardy section of the Italian Society of Infectious and Tropical Diseases (SIMIT), proposed the lopinavir/ritonavir (LPV/r) and hydroxychloroquine (HCQ) combination as a therapeutic protocol for hospitalised patients with the respiratory symptoms associated with COVID-19 [6, 7]. This indication was based on experimental studies showing that HCQ (an antimalarial drug that is also widely used to treat autoimmune disorders) has *in vitro* antiviral activity of against SARS-CoV-1, human coronavirus 229E (HCoV-229E) and SARS-CoV-2 [8–10], and it has been postulated that it may benefit patients with COVID-19 because of its modulatory effects on the production and release of tumor necrosis factor 1 (TNF-1) and interleukin-6 (IL-6), both of which are thought to be involved in the inflammatory damage associated with late-stage COVID-19 [10, 11]. There were also data indicating that LPV, an HIV-1 aspartate protease, has *in vitro* activity against SARS-CoV-1 and MERS coronavirus (MERS-CoV) [12, 13], and a clinical study conducted in Hong Kong in 2003 found that the addition of LPV co-formulated with ritonavir (LPV/r) to a standard treatment protocol (ribavirin plus steroid therapy) was associated with improved clinical outcomes of patients affected by SARS-CoV-1 [14].

However, very recent studies have questioned the clinical efficacy of LPV/r and HCQ against COVID-19. In particular, one randomised controlled trial comparing the efficacy of LPV/r with that of standard of care in patients with severe COVID-19 did not find any significant differences in mortality, clinical improvement or viral shedding [15], and an observational study carried out in New York did not find any difference in mortality between severely ill patients with COVID-19 who received HCQ and those who did not [16]. However, neither of these studies considered the possible effect of the timing of the start of treatment, although there is evidence that early treatment is crucial when assessing efficacy against acute respiratory infections [17–20].

The aim of this study was to analyse the combined effect of LPV/r and HCQ treatment on the course of COVID-19 by examining differences in the clinical outcomes of patients who started treatment within five days of the onset of symptoms and those who started later.

## 2. PATIENTS AND METHODS

This retrospective cohort study involved patients with COVID-19 pneumonia who were hospitalised at Luigi Sacco Hospital, Milan, Italy, between 21 February and 20 March 2020. COVID-19 pneumonia was diagnosed on the basis of the detection of SARS-CoV-2 RNA on nasopharyngeal swab using a real-time reverse-transcriptase polymerase chain reaction (RT-PCR) test processed using the automated ELITe InGenius® system and the GeneFinder™ COVID-19 Plus RealAmp Kit assay (ELITechGroup, Puteaux, France) and a chest X-ray with signs of pneumonia or ≤ 93% oxygen saturation (SpO_2_) while breathing room air [21].

In accordance with the SIMIT drug protocol, all patients with COVID-19 pneumonia admitted to our hospital during the study period were offered off-label treatment with LPV/r 400/100 mg (tablet COVID-19 Plus or oral solution) twice daily plus hydroxychloroquine 200 mg twice daily for a minimum of five and a maximum of 20 days depending on patients’ clinical response [6, 7]. The exclusion criteria were the presence of any condition that would not allow the treatment to be safely administered (including any known allergy or hypersensitivity to the drugs used in the protocol); severe liver or kidney disease; the use of medications contraindicated with LPV/r that could not be replaced or discontinued; pregnancy or breast-feeding; known HIV infection; a history of cardiomyopathy, arrhythmias or conduction disorders; and a history of ocular macular disease or retinal damage.

The patients were included in the intention-to-treat analysis if they had received at least one dose of the scheduled treatment. Patients who died on the day of starting treatment were excluded from the analysis.

The study was approved by hospital’s ethical committee (*Comitato Etico Interaziendale Area 1*), and all of the study patients gave their written informed consent to the administration of off-label treatment (informed consent was waived in the case of those undergoing mechanical ventilation).

### 2.1 Data collection

The collected data included demographic data, the Charlson Comorbidity Index (CCI) unadjusted for age, date of onset of symptoms, signs and symptoms at the time of presentation, laboratory findings, and disease severity at the time of starting the study treatment. In accordance with the China Guidelines for the Diagnosis and Treatment of Novel Coronavirus (2019-nCoV) Infection, severity was classified as mild (only slight clinical symptoms and no imaging of pneumonia), moderate (with fever, respiratory symptoms and confirmed pneumonia), severe (with respiratory distress [>30 breaths per minute], or < 93% resting oxygen saturation or PaO_2_/FiO_2_ < 300 mmHg), or critically severe (with respiratory failure requiring mechanical ventilation, or shock, or any other organ failure needing intensive care) [22].

The patients’ clinical status was monitored from the day of treatment initiation to day 30, and data concerning the requirement of oxygen support, laboratory values, serious adverse events, and discharge or death were recorded. The living status of the patients discharged before day 30 was assessed by means of telephone calls to the patients themselves.

### 2.2 Outcomes

The primary outcome was clinical improvement, defined as a decrease from baseline of at least two categories of the seven-category ordinal scale recommended by the WHO R&D Blueprint Group [23], which consists of 1 = not hospitalised, capable of resuming normal activities; 2 = not hospitalised, but unable to resume normal activities; 3 = hospitalised, but not requiring oxygen supplementation; 4 = hospitalised and requiring oxygen therapy; 5 = hospitalised and requiring high-flow nasal oxygen therapy, non-invasive mechanical ventilation, or both; 6 = intensive care unit (ICU) hospitalisation, requiring invasive mechanical ventilation or extra corporeal membrane oxygenation (ECMO), or both; 7 = deceased.

The secondary outcomes were 30-day mortality and drug safety, including adverse events leading to premature treatment discontinuation. Adverse events were classified using the National Cancer Institute Common Terminology Criteria for Adverse Events, version 4.0.

### 2.3 Statistical analysis

The study population was divided into two groups: an early treatment (ET) group of patients who started LPV+HCQ treatment < 5 days from the onset of symptoms; and a delayed treatment (DT) group of patients who started treatment ≥ 5 days from the onset of symptoms.

The baseline demographic and clinical characteristics of the two groups were compared using the χ^2^ (or Fisher’s exact test where necessary) for categorical variables, and Wilcoxon’s rank-sum test for continuous variables. The cumulative incidence of clinical improvement from day 1 (treatment start) to day 30 was estimated using death as a competing event and compared between groups using Gray’s test. Uni- and multivariable logistic regression models were used to assess the influence of the timing of the start of treatment on the probability of 30-day mortality. All of the factors judged to be clinically relevant to the study outcome were considered possible confounders in the multivariable model. The data were analysed using SAS software, version 9.4, and a p-value of < 0.05 was considered statistically significant.

## 3. RESULTS

Between 21 February and 20 March 2020, 172 patients with COVID-19 pneumonia started LPV+HCQ treatment at our Hospital and received at least one dose: 43 (25%) in the ET group and 129 (75%) in the DT group. The median time from the onset of symptoms to starting the study treatment was three days (interquartile range [IQR] 2.5-4) in the ET group and eight days (IQR 6-10) in the DT group. The majority of the patients were males (72.1%) in their sixties presenting with moderate (53.4%) or severe disease (34.9%) associated with fever (72.7%).

Table 1 shows the baseline clinical and laboratory characteristics of the patients in the two groups. There were no significant between-groups differences in terms of their demographic characteristics or disease severity, but the patients in the DT group had a higher burden of co-morbidities (median CCI = 3, IQR 1-5 *vs* 2, IQR 0-3; *p* = 0.041), and more frequently presented with cough (58.9% *vs* 39.5%; *p* = 0.034) and fever (76.7% *vs* 60.4%; *p* = 0.045). They also had higher median white blood cell (*p* = 0.017) and neutrophil counts (*p =* 0.030), higher median C-reactive protein levels (*p* = 0.045) and lower median PaO_2_ levels (*p* < 0.001).

The median duration of LPV/r+HCQ treatment was six days (IQR 5-8), with no significant difference between the groups.

**Table 1.**
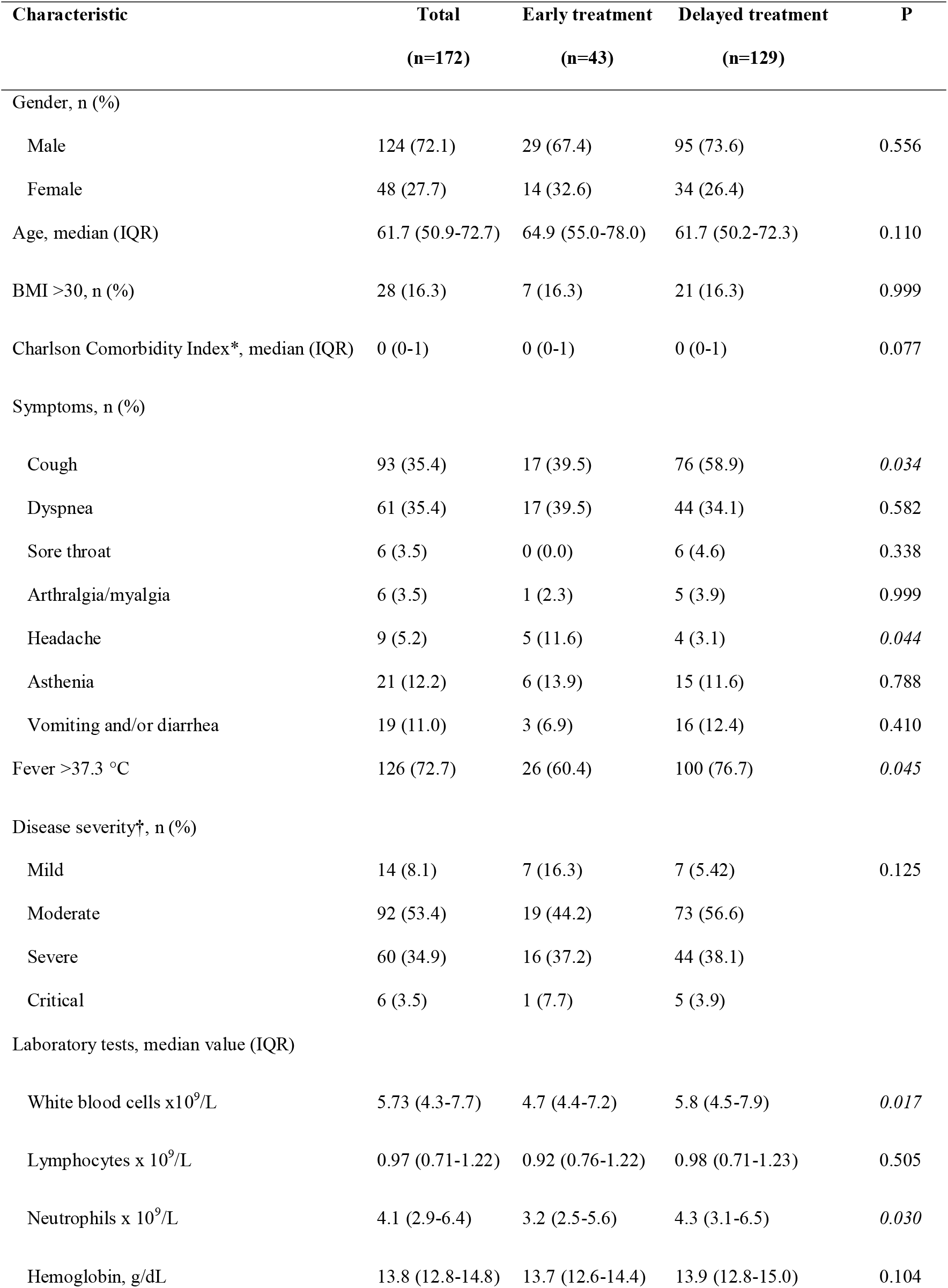
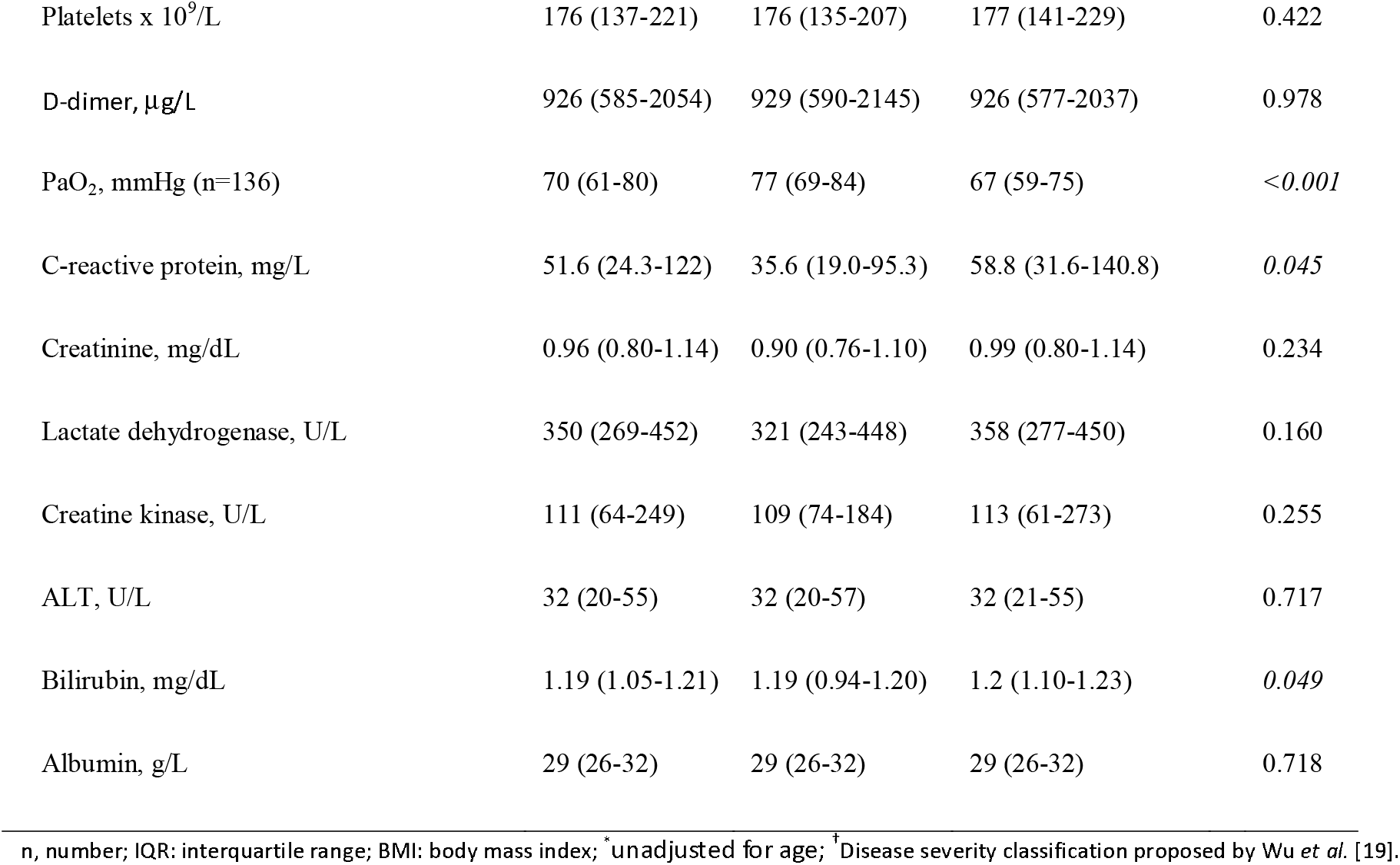
Baseline characteristics of the study population at LPV/r+HCQ initiation.

Forty patients (22.7%) discontinued the treatment before completing the minimum 5-day course, with no significant difference between the ET and DT group (16.3% *vs* 25.5%; *p* = 0.296). The reasons for discontinuing were a switch to another treatment protocol (18, 45%), adverse events (n = 14, 35%), early discharge (n = 5, 12.5%), death (n = 2, 5%), and possible interaction with other treatments (n = 1).

Sixty patients (34.9%: 19 [11.0%] who prematurely discontinued LPV/r+HCQ treatment and 41 [23.8%] who received it for > 5 days) were administered other treatment/s during the study period, including remdesivir (n = 33, 19.2%), tocilizumab (n = 36, 20.9%) or both (n = 10, 5.8%). The proportion of patients who received other treatments was not significantly different between the two groups: remdesivir was given to four ET patients (9.1%) and 29 DT patients (22.5%) (*p* = 0.057), and tocilizumab was given to respectively six (13.6%) and 30 patients (23.5%) (*p* = 0.193).

### 3.1 Treatment outcomes

As shown in Figure 1, the cumulative incidence of clinical improvement increased over time from 36.6% on day 10 to 66.3% on day 20 and 73.3% on day 30, with no significant difference between the two groups (*p* = 0.213) (Fig. 2).

**Figure 1.**
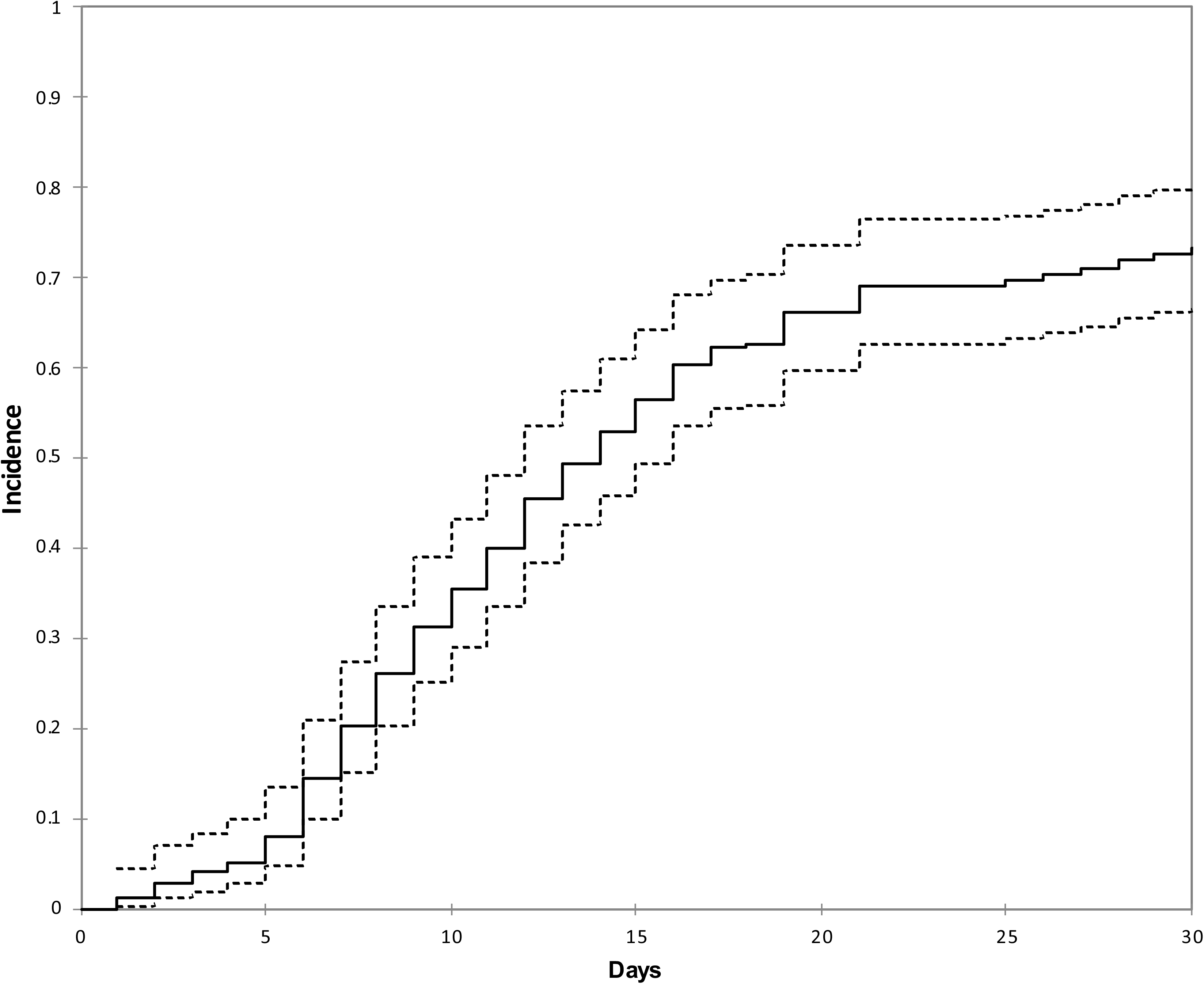
Cumulative incidence of improvement (solid line) and 95%Cis (dashed lines).

**Figure 2.**
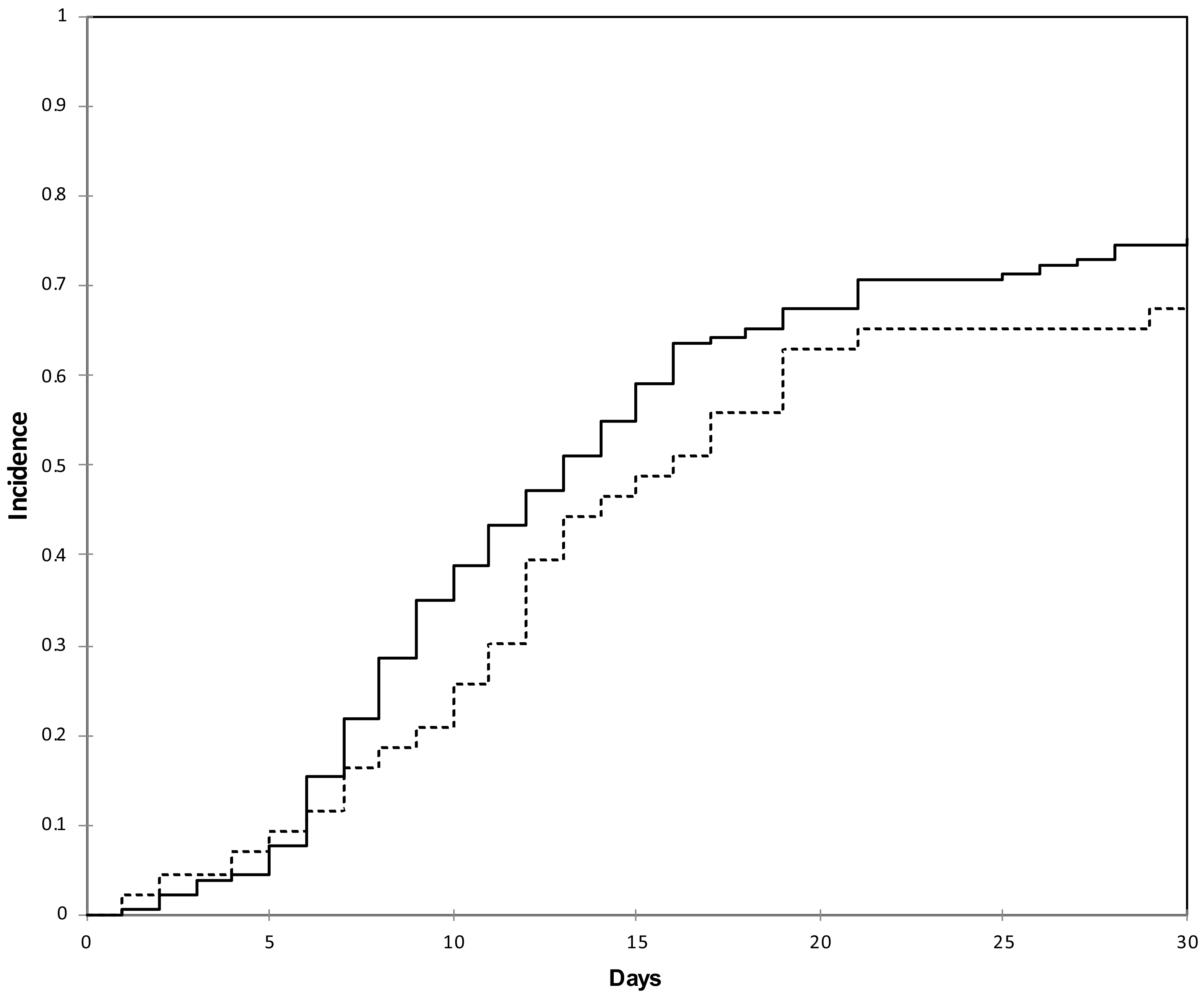
Cumulative incidence of improvement in the ET group (dashed line) vs DT group (solid line).

At the end of the study period, 23.2% of the patients in the ET group and 17% of those in the DT group had died. The univariable analysis did not reveal any significant association between the timing of the start of LPV/r+HCQ treatment and the probability of 30-day mortality (odds ratio [OR] of < 5 days *vs* ≥ 5 days 1.58, 95% confidence interval [CI] 0.70-3.56; *p* = 0.271). After adjusting for relevant clinical variables in the multivariable model, an earlier start of treatment was still not associated with a lower probability of 30-day mortality (adjusted OR [aOR] of < 5 days *vs* ≥ 5 days 1.45, 95%CI 0.50-4.19) (Fig. 3). Conversely, age per ten years more (aOR 2.21, 95%CI 1.38-3.57), obesity (aOR 3.90, 95%CI 1.19-12.82), and undergoing invasive or non-invasive mechanical ventilation (aOR 4.75, 95%CI 1.38-16.34) were all independently associated with an increased probability of death (Fig. 3).

**Figure 3.**
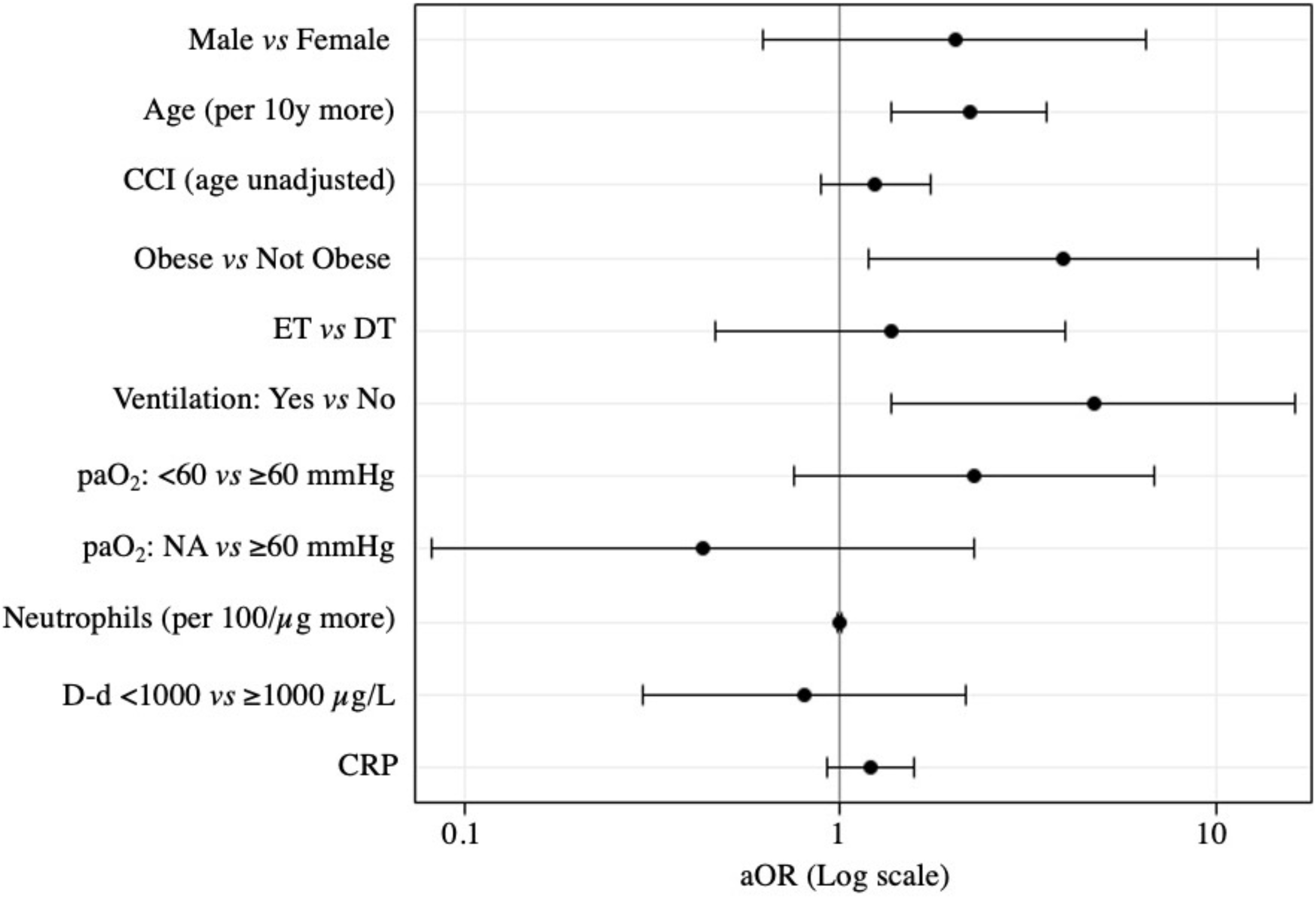
Multivariable model results (adjusted odds ratios).

### 3.2 Safety

The most frequent adverse events were an increase in hepatic enzymes to at least five times above the normal values (13 patients, 7.6 %), and grade 2-3 nausea and/or diarrhoea (14 patients, 8.1%). The treatment was discontinued in all of the 14 patients who developed grade 2-3 gastrointestinal disorders.

## 4. DISCUSSION

The spread of the COVID-19 pandemic and the exponential increase in deaths worldwide has made the demand for clinical evidence concerning new and pre-existing drugs increasingly pressing. Various molecules, including antivirals and immune modifiers, were rapidly evaluated in initial uncontrolled studies and are now being investigated in randomised controlled trials.

The search for an effective treatment of COVID-19 also needs to consider the optimal time to start the use of effective drugs, taking advantage of the emerging data concerning the pathogenetic mechanisms underlying different stages of the disease. As it has been shown that the pathogenesis of COVID-19 includes a viremic phase that peaks 5-6 days after infection, followed by an immune-mediated phase characterised by an aggressive inflammatory response that is largely responsible for airway damage [4], it is possible to hypothesise that the early use of effective antiviral drugs would reduce the progression and mortality of COVID-19, as has been observed in the case of other acute viral respiratory illnesses [17–20].

However, our study assessing possible differences in the clinical outcomes of patients who received LPV/r+HCQ < 5 or > 5 days after symptom onset did not reveal any difference in the time to clinical improvement or in the probability of 30-day mortality between the two groups. This raises some doubts about the *in vivo* effect of LPV/r+HCQ treatment on SARS-CoV-2, which are also supported by emerging pharmacological questions. It has been recently estimated that the protein-adjusted 90% inhibitory concentrations (PA-IC_90_) of LPV required to inhibit SARS-CoV-2 replication in plasma, epithelial lining fluid (ELF) and cerebrospinal fluid (CSF) are respectively 200-fold, 20-fold and 2000-fold higher than those measured *in vivo* [24]. Moreover, a recently published mechanistic model has shown that, instead of the conventional lower dose of ≤ 400 mg/day, HCQ doses of > 400 mg twice daily for ≥ 5 days would be required to obtain a rapid decrease in viral load, a reduction in the proportion of patients with detectable SARS-CoV-2 infection, and shorter treatment courses [25], but it has been predicted that doses of > 600 mg twice daily would prolong the QT interval and lead to a risk of arrhythmias, including *torsade de pointes* [25].

A total of 14 (8.1%) patients in our study were unable to complete the minimum 5-day course of LPV/r+HCQ because of adverse events. The most frequent severe adverse events were gastrointestinal disorders (nausea and/or diarrhoea) mainly attributed to LPV/r. Interestingly, a recent study has found that the trough concentrations of LPV measured in COVID-19 patients are three times higher than those measured in HIV patients, which may explain why COVID-19 patients poorly tolerate LPV [26]. Furthermore, Cao *et al*. found that nearly 14% of the patients who received LPV/r in their randomised trial could not complete the full course of 14 days mainly because of gastrointestinal intolerance [15] and, as they did not find that LPV/r had a beneficial effect on the clinical course of COVID-19, they suggest that its use may expose COVID-19 patients to unnecessary toxicities.

Our study has a number of limitations. Firstly, given the emergency context in which it was carried out, it was impossible to include a control group, and so we cannot exclude the possibility that the patients whose status improved after LPV/r+HCQ treatment would have improved regardless of any treatment. Secondly, a relatively large proportion of our patients received other experimental treatments during the study period, and this is clearly a confounding factor when analysing the efficacy LPV/r+HCQ: however, as there was no between-group difference in the proportion of patients who received other treatments, it is likely that this had no impact on our analysis of the effect that the time of starting treatment had on COVID-19 outcomes.

Thirdly, the treatment’s virological efficacy (i.e. the reduction in viral load in nasopharyngeal secretions) could not be assessed because there was no regular monitoring of the presence of SARSCoV-2 genome on nasopharyngeal swabs and the RT-PCR available in our microbiology department only provides qualitative data.

Finally, the study was conducted in the ever-changing scenario created by the dramatic escalation of the epidemic in Northern Italy. The Infectious Diseases Department of Luigi Sacco Hospital acts as a north Italian reference centre for infectious diseases. Consequently, our findings concerning the potential use of LPV/r+HCQ relate to hospitalised patients in the early wave of the Italian pandemic and may not extend inferred to outpatients with milder symptoms.

In conclusion, we found that starting LPV/r+HCQ treatment within five days of symptom onset was not associated with a more rapid improvement in the clinical condition of patients hospitalized with COVID-19 or a reduced probability of 30-day mortality. Together with the relatively high rate of severe adverse event attributable to LPV/r, this raises some doubts about the benefit of combined LPV/r and HCQ treatment of COVID-19. More rigorous controlled studies are needed to assess the real benefit-to-harm ratio of LPV/r and HCQ, and the use of the combination should be discouraged in other contexts.

## Data Availability

The datasets used and/or analysed during the current study are available from the corresponding author on reasonable request.

CCI: Charlson Comorbidity Index
ET: early treatment group
DT: delayed treatment group
paO_2_: partial oxygen pressure
D-d: D-dimer
NA: not assigned
CRP: C-reactive protein
aOR: adjusted odds ratio
Log: logarithmic.

## LIST OF ABBREVIATIONS

COVID-19: coronavirus induced disease
SARS-CoV-2: severe acute respiratory syndrome coronavirus 2
ET: early treatment
DT: delayed treatment
HCoV-229E: Human coronavirus 229E
TNF-1: Tumor necrosis factor 1
IL-6: interleukin 6
MERS: Middle Eastern Respiratory Syndrome Coronavirus
LPV/r: lopinavir/ritonavir
HCQ: hydroxychloroquine
SIMIT: Società Italina di Malattie Infettive e Tropicali
RT-PCR: real time polymerase chain reaction
SpO2: percutaneous oxygen saturation
CCI: Charlson comorbidity index
FiO2: fraction of inspired oxygen
PaO2: arterial partial pressure of oxygen
IQR: inter quartile range
ICU: intensive care unit
ECMO: Extra corporeal membrane oxygenation
CRP: C-reactive protein
OR: Odds Ratio
aOR: adjusted Odds Ratio
CI: Confidence Interval
AE: Adverse Events
PA-IC90: protein-adjusted 90% inhibitory concentration
ELF: epithelial lining fluid
CSF: cerebrospinal fluid

## Authors’ contributions

AG, GP, ALR and MG designed the study. LO, AG, GP and ALR were responsible for the statistical analysis. All authors contributed to patient enrolment, and data collection and interpretation. AG and GP drafted the manuscript, which was critically reviewed by ALR, MG, SA, SR. All authors approved the final version of the manuscript.

## Availability of data and materials

The datasets used during the current study are available from the corresponding author upon reasonable request.

## Acknowledgements

The authors would like to thank all of the patients and their families and all of the medical staff (paramedics, nurses and physicians) who are making every effort to ensure the best care for patients suffering from coronavirus disease.

---

**Table 1.**
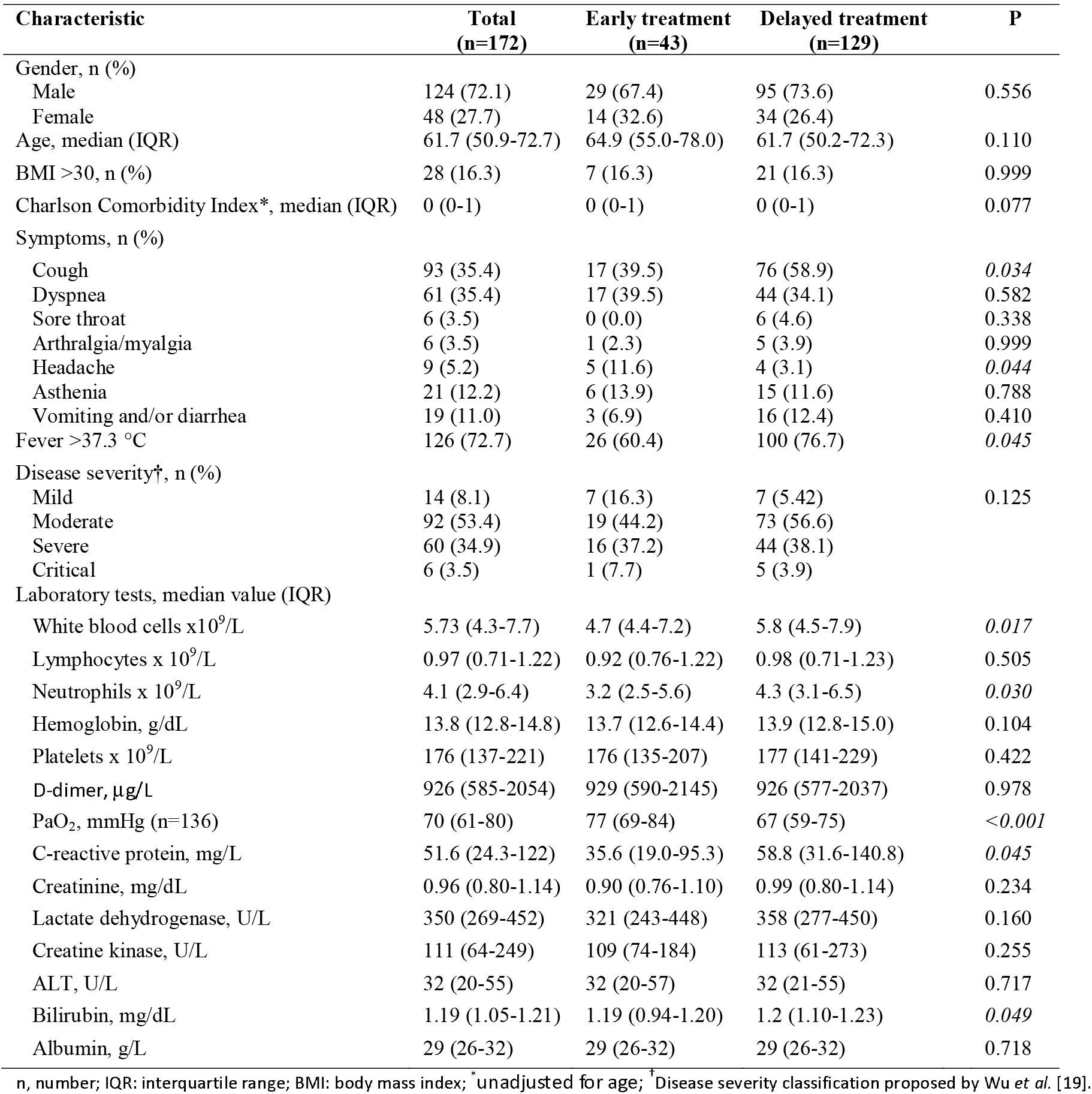
Baseline characteristics of the study population at LPV/r+HCQ initiation Characteristic Total (n = 172)

